# Tuberculous meningitis in adult patients of a Mexican neurological referral center: A-six-year retrospective cohort study

**DOI:** 10.1101/2020.12.02.20242420

**Authors:** Maldonado-Diaz Ellis Daniela, Soto-Hernández José Luis, Salinas-Lara Citlaltepetl, Kammar-Garcia Ashuin, Cárdenas Graciela

## Abstract

**Introduction:** Tuberculosis (TB) remains as an important concern of public health worldwide because the high prevalence and severe sequelae. Tuberculous meningitis (TBM) is the most lethal and disabling form.

**Aim:** To describe the clinical, laboratory, and neuroimaging characteristics of TBM on admission at neurological center in Mexico City.

**Methods:** Retrospective cohort study at the third level neurological center from 2010 to 2016. Clinical follow-up was evaluated at hospital discharge, three months, and one-year either due to lack of follow-up or mortality, during the follow-up the adverse events were registered.

**Results:** One-hundred and six patients were included, 74 (69.8%) males and 32 (30.2%) females. From these 31 (29.2%) were HIV-positive. The median age was 35.5 (IQR:28-51). Pulmonary TB was found in 25% of the population. Alcoholism was observed in a half of the patients while diabetes in 15%, the latter being significant (p=0.04). Abnormalities in neuroimaging were significant among our population (p=0.003). Only one-third of the population had a positive Lowenstein-Jensen culture. There were no differences in clinical outcomes between HIV positive and non-HIV patients.

**Conclusion:** Our study shows data to those described in the literature. The initiation of empirical treatment in all patients with a high clinical suspicion of tuberculosis mandatory to try to avoid severe neurological sequels.

## Introduction

Tuberculous meningitis (TBM) is the most frequent form of central nervous system (CNS) tuberculosis (1). It represents 1% per year of all tuberculosis (TB) cases, highly disabling and even lethal (1,2).

Globally, TB remains, a chief health problem with an estimated 10.0 million cases and 1.2 million deaths resulting from disease during 2018 (WHO report, 2019), but the global TB burden remains unclear because of underreporting and underdiagnosing. (3). TB is endemic in Mexico, which has the third-highest prevalence in Latin America, just below Brazil and Peru. During 2019, there were 45,637 cases of pulmonary TB and 396 cases of TBM (4). Many factors are related particularly the high prevalence of risk population as diabetes (DM), HIV/AIDS, and poverty. HIV infection represents the highest risk for latent infection and the progress of clinical TB (5). Other immunocompromised states like chronic steroid use and chronic alcoholism carry the same risk for developing TBM (6,7).

This manuscript aims to describe the clinical, laboratory, and neuroimaging characteristics of TBM patients admitted at neurological center in Mexico City.

## Material y methods

### Study design and patient selection

Retrospective cohort study carried out in a third level neurological center (Instituto Nacional de Neurología y Neurocirugía “Manuel Velasco Suarez”). The diagnosis was based on Marais Consensus of TBM (8). Only patients with definitive and probable TBM diagnosis were included.

The Modified British Medical Research Council criteria was used to define TBM severity (9), and the comorbidity was defined by Charlson Comorbidity Index (CCI) (10). Besides, a nutritional assessment was performed by the body mass index (BMI) in six categories, as follows ≤18.5 malnourishment, 18.5-24.9 average weight, overweight ≥ 25, and obesity ≥ 30 (11). Strictly neurological examinations were performed to state sequels.

Clinical follow-up was evaluated at hospital discharge, three-months, and one-year after hospitalization, either due to lack of follow-up or mortality. All adverse events were register during the clinical follow-up.

This study protocol was approved (protocol 85/17) by the research and ethics committees of the Instituto Nacional de Neurología y Neurocirugía “Manuel Velasco Suárez”.

### Statistical analysis

All qualitative variables were expressed in frequencies and percentages. The data were compared between HIV-positive and Non-HIV patients; the categorical comparison was made using the χ^*2*^ or Fisher’s exact test. Quantitative variables were expressed as median and rage. A Mann-Whitney test based on data behavior in the Kolmogorov-Smirnov test was used for comparison between groups. Differences were considered, statistically significant at p<0.05. Data were analyzed using SPSS v.21 (IBM Inc., Armonk, NY).

## Results

### Epidemiological data

From January 2010 to December 2016, One-hundred and six patients and the clinical, laboratory and neuroimaging characteristics were collected from medical records. For statistical analysis, two groups were considered: HIV-positive 31 (29 %) and non-HIV infected 75 (71%). In the first group, the median viral load was 146657 copies (IQR: 73006.5-555664.8), median T CD4 lymphocyte cell count of 10 (IQR: 3.3-16.5), and median T CD8 lymphocyte cell count of 63 (IQR 52-78). Male population predominated 74 (70 %) (p=0.003).

### Clinical and neuroimaging characteristics

The median of age was 35.5 (IQR:28-51), higher in non-HIV infected. The weight and body mass index was similar between the HIV infected and non-infected patients. Several risk factors were observed, alcoholism in 55 (52 %), diabetes mellitus in 16 (15%); interestingly, previous pulmonary TB was observed in 24 (25%). Only diabetes was statistically different between groups, predominated in non-HIV patients (p=0.04) (Table 1).

**Table 1.** Demographic characteristics of patients

|  | **Total**<br>**n=106** | **HIV**<br>**infected**<br>**n=31** | **Non-HIV**<br>**infected**<br>**n=75** | **p value** |
| --- | --- | --- | --- | --- |
| Sex, n (%) |  |  |  |  |
| Woman | 32 (30.2) | 3 (9.7) | 29 (38.7) | 0.003 |
| Men | 74 (69.8) | 28 (90.3) | 46 (61.3) |  |
| Weight, Kg | 70 (60-77.4) | 70 (62.9-79) | 70 (59-77) | 0.4 |
| Body mass index | 25 (23.4-28) | 24.5 (21.7-27.8) | 25 (23.5-28.1) | 0.4 |
| Age, years | 35.5 (28-51) | 32 (29-35) | 42 (28-55) | 0.01 |
| Age groups, n (%) |  |  |  |  |
| ≤20 | 9 (8.5) | 1 (3.2) | 8 (10.7) | 0.02 |
| 21-30 | 27 (25.5) | 11 (35.5) | 16 (21.3) |  |
| 31-40 | 26 (24.5) | 13 (41.9) | 13 (17.3) |  |
| 41-50 | 15 (14.2) | 4 (12.9) | 11 (14.7) |  |
| 50-59 | 16 (15.1) | 2 (6.5) | 14 (18.7) |  |
| >60 | 13 (12.3) | 0 (0) | 13 (17.3) |  |
| Risk factors, n (%) |  |  |  |  |
| Previously lung TB | 24 (25) | 8 (28.6) | 16 (23.5) | 0.6 |
| DM | 16 (15.1) | 1 (3.2) | 15 (20) | 0.04 |
| Malnourishment | 13 (12.6) | 4 (12.9) | 9 (12.5) | 0.9 |
| Alcoholism | 55 (51.9.) | 20 (64.5) | 35 (46.7) | 0.09 |
| Cocain | 3 (2.8) | 2 (6.5) | 1 (1.3) | 0.9 |
| Marihuana | 7 (6.6) | 4 (12.9) | 3 (4) | 0.9 |
| Methamphetamines | 1 (0.9) | 1 (3.2) | 0 (0) | 0.9 |
| Data are presented as median (IQR) and n (%). Data were compared by Man-Whitney U test and Chi Square or Fisher exact test. |  |  |  |  |

Clinical symptoms included systemic one (weight loss 38% and fever 50%), neurological symptoms (dizziness, lethargy, or altered consciousness) observed in 80 patients (76%), include seizures in 70 (66%) and meningeal sign in 49 (46%). No statistical differences were observed between groups (Table 2).

**Table 2.** Clinical characteristics of population

|  | Total<br>n=106 | HIV<br>infected<br>n=31 | Non-HIV<br>infected<br>n=75 | p value |
| --- | --- | --- | --- | --- |
| Weight loss, n (%) | 40 (37.7) | 14 (45.2) | 26 (34.7) | 0.4 |
| Fever, n (%) | 52 (49.5) | 15 (50) | 37 (49.3) | 0.9 |
| Combe test, n (%) | 20 (18.9) | 6 (19.4) | 14 (18.7) | 0.9 |
| Meningean signs, n (%) | 49 (46.2) | 12 (38.7) | 37 (49.3) | 0.3 |
| Seizures, n (%) | 70 (66) | 21 (67.7) | 49 (65.3) | 0.8 |
| Neurological symptoms,<br>n (%) | 80 (75.5) | 24 (77.4) | 56 (74.6) | 0.6 |
| GCS | 15 (15-15) | 15 (15-15) | 15 (15-15) | 0.3 |
| Severity, n (%) |  |  |  |  |
| BRMC I | 51 (48.1) | 9 (29) | 42 (56) | 0.09 |
| BRMC II | 38 (35.8) | 17 (54.8) | 21 (28) |  |
| BRMC III | 17 (16) | 5 (16.1) | 12 (16) |  |
| CCI | 1 (0-6) | 6 (6-7) | 2 (1-3) | <0.0001 |
| Comorbidity, n (%) |  |  |  |  |
| Absent | 58 (54.7) | 2 (6.5) | 56 (74.7) | <0.0001 |
| Low | 9 (8.5) | 1 (3.2) | 8 (10.7) |  |
| Hight | 39 (36.8) | 28 (90.3) | 11 (14.7) |  |
| Nutritional status, n (%) | 106 | 31 | 75 |  |
| Malnourishment | 4 (3.8) | 2 (6.5) | 2 (2.6) | 0.4 |
| Normal weight | 49 (46.2) | 17 (54.8) | 32 (42.7) |  |
| Overweight | 39 (36.9) | 11 (35.5) | 28 (37.3) |  |
| Obesity I | 10 (9.4) | 1 (3.2) | 9 (12) |  |
| Obesity II | 3 (2.8) | 2 (6.5) | 1 (1.3) |  |
| Obesity III | 1 (0.9) | 0 (0) | 1 (1.3) |  |
| Data are presented as median (IQR) or n (%). Data were compared by Man-Whitney U test and Chi Square or Fisher exact test.<br>GCS: Glasgow coma scale, CCI: Charlson comorbidity index |  |  |  |  |

According to the British Medical Research Council, stage I, was observed in 51 patients (48%), grade II in 38 patients (36%), and finally, grade III in 17 patients (16%); no statistical differences were observed between HIV-positive and non-HIV infected.

Charlson’s Comorbidity Index was also evaluated. Low risk was observed in 9% (9) and high risk in 37% (39) as high risk. In this latter, a significantly higher proportion of patients were HIV-positive (p=0.0001).

### CSF cytochemical and laboratory findings

All patients showed inflammatory profiles in the cytochemical analysis of CSF. Median glycorrhachia was 47 mg/dL (ranges 18-64), proteinorachia 95 mg/dL (44-188) and cellularity 53.5 cells/mm^3^ (6-195). Th median CSF adenosine deaminase level was 7 UI/L (3-14). No differences were observed between groups. Median leukocytes were 8.300 mm^3^ (5.9-10.7). Non-HIV infected showed leukocytosis (p=0.001). Only one-third of the patients had any Lowenstein-Jensen cultures (Table 3).

**Table 3.** Ancillary test performed in patients

|  | **Total**<br>**n=106** | **HIV positive**<br>**n=31** | **Non-HIV**<br>**n=75** | **p value** |
| --- | --- | --- | --- | --- |
| Serum |  |  |  |  |
| Leukocytes, x10<sup>9</sup>/L | 8.3 (5.9-10.7) | 6.8 (5.4-8.9) | 8.9 (6.7-11.8) | 0.001 |
| Sodium, mmol/L | 136 (133-139) | 136 (134-141) | 136 (133-139) | 0.5 |
| Glucose, mg/dL | 106 (89-125.5) | 96 (88-113) | 109 (91-180.5) | 0.1 |
| CSF |  |  |  |  |
| Glucose, mg/dL | 47 (18.3-63.8) | 44 (34.5-61.5) | 48 (16.8-65.3) | 0.7 |
| Proteins, mg/dL | 95 (44-188) | 98 (44.5-200) | 92 (43-186.5) | 0.9 |
| Cells | 53.5 (6-195) | 49.5 (7-141) | 63 (5.8-213) | 0.7 |
| Lymphocytes | 90 (66.3-97) | 90.5 (66-96.5) | 87 (66.3-97.8) | 0.8 |
| Neutrophils | 14 (4.5-38.5) | 10 (5-83) | 15.5 (3.8-35.8) | 0.5 |
| Adenosine deaminase, UI/L | 7 (3-14) | 7 (3.8-13) | 6 (1.5-15) | 0.7 |
| Microbiological test |  |  |  |  |
| Mtb positive | 33 (31.1) | 10 (32.3) | 23 (30.7) | 0.9 |
| Data are presented as median (IQR) Data were compared by Man-Whitney U test.<br>CSF: Cerebro spinal fluid, Mtb: Mycobacterium tuberculosis |  |  |  |  |

Neuroimaging was performed in 79 (75%) of patients during hospitalization: Main abnormalities which predominated in non-HIV infected were vasculitis (p=0.03), parenchymal granulomas (p=0.02) and basal enhancement (p=0.06), no differences between groups were observed in hydrocephalus (Table 4). Transcranial doppler showed vasculitis in 37.7% of patients.

**Table 4.** Neuroimaging characteristics

|  | **Total**<br>**n=106** | **HIV positive**<br>**n=31** | **Non-HIV**<br>**infected**<br>**n=75** | **p value** |
| --- | --- | --- | --- | --- |
| Vasculitis | 37 (34.9) | 6 (19.4) | 31 (41.3) | 0.03 |
| Hydrocephalus | 9 (8.5) | 1 (3.2) | 8 (10.7) | 0.2 |
| Meningitis | 42 (36.6) | 8 (25.8) | 34 (45.3) | 0.06 |
| Tuberculoma | 57 (53.8) | 11 (35.4) | 46 (61.3) | 0.02 |
| Data are presented as n (%). Data were compared Chi Square or Fisher exact test. |  |  |  |  |

### Treatment

All patients received antituberculous (DOTBAL) treatment a combination of Ethambutol, Pyrazinamide, Isoniazid, and rifampicin (Intensive phase) for two months and Isoniazid and rifampicin (Support phase) for 12 to 15 months. During hospitalization, patients also received intravenous dexamethasone at 0.3mg/kg/day for two weeks and later, switched on oral prednisone with gradually tapered. Antiretroviral therapy (ART) was started in the population with HIV-positive; median viral load at six months after treatment was 131 (IQR: 40-278), CD4 cell count 170 (IQR: 149-279), and CD8 cell count 55.5 (IQR: 43.5-62.3). There were no significant differences between survivors and those who died during the study.

### Neurological sequels and clinical outcomes

Surviving patients were 86 (81%), and 49 (57%) developed neurological sequels, particularly motor deficits in 23%, cranial neuropathy in 20% and cognitive complaints in 14%. The whole mortality rate was 19%, without differences between groups. In HIV-positive patient, mortality was more frequently (19.4% vs. 18.7%) observed during hospitalization while non-HIV infected, 85% died within the first year of clinical follow-up.

## Discussion

Meningeal tuberculosis is one of the most difficult causes of meningitis to diagnose due to the difficulties in identifying mycobacteria, which is why any patient with any symptoms and signs of meningitis should be considered TB infected in regions with a high prevalence or in population of high risk. It is important to state that Mexico has the third place of TB prevalence after Brazil and Peru (WHO 2019), TBM is an infectious disease with high rates of disability and mortality (12), worldwide the prevalence of disseminated disease vary between 30-50% of infected patients (WHO 2019) while in the current report, our series showed 25% of active lung TB.

According to TB risk factors, HIV and diabetes are the main described in the literature (13) but others such as drugs abuse, and alcoholism are important. In a Mexican cohort study in Tijuana, Baja California includes 41 TB patients, in which 51% were drug users, and 27% of alcoholics (14). This data contrasted with our series where only 10% were drug users while 52% alcoholics.

Clinical presentation of TBM is highly pleomorphic but includes cranial nerve palsy, headache, meningeal syndrome, and mental alterations. A study carried out in Brazil in 2013, in which 300 patients were included, reported upon admission alterations in the state of consciousness in 61% (15) while in our series 16% of population had coma, this data may indicate a delay in the diagnosis due to delays in consultation.

Delays in the diagnosis seem to be a common problem in low-income countries, in fact in a Peruvian study of 3802 patients, the clinical severity according to British Medical Research in most than a half of population was stage II (16), another example is the prospective cohort conducted by Siddiqi Omar in Zambia (Siddiqi OK et al. 2019) with 500 TB patients were almost a half (46%) presented stage II. In comparison in our series, 48% presented stage I.

Neuroimaging is an important diagnostic tool, particularly in CNS TB where bacteriological or molecular diagnosis (geneXpert) is less sensitive (17). It is frequent to found neuroimaging abnormalities in patients with severe disease (18). In our institution, the first performed approach was CT-scan, where specific findings as vasculitis, hydrocephalus and granulomas are highly suspicious of TBM. CT-scan abnormalities were, frequent in non-HIV infected. In our series, it was observed on Transcranial doppler an increased flow velocity in the middle cerebral artery (MCA) in 38%, this data contrast with a report in a TBM pediatric population where up to 70% of the population presented vasculitis (19). In a Malaysian study, in a cohort of 51 patients it was observed cerebral infarcts in 67% (the most frequently affected areas were basal ganglia) as well as secondary vasculitis in MCA in 25% of population (20).

The global mortality reported in our series was 19%, which is comparable with another Mexican study data conducted by García-Grimshaw M. Et al. In this series a cohort of 41 adult patients was reported. The median age was 28-years old and mortality of 19.5% (14).

TBM has been reported to enhance HIV replication in alveolar macrophages and peripheral blood T cells through cytokine and antigen-mediated cell activation. In a study carried out in South Africa (21), 105 patients were included, all HIV-positive without antiretroviral therapy, reported a T CD4 lymphocytes count upon admission of 78 (IQR: 45-286) while after the initiation of ARV treatment the T CD4 lymphocytes count was 130 (IQR: 76-336) with no statistically significant difference, our population had a similar behavior.

In a Vietnam cohort study of 1699 TBM patients, it was reported that 951 were non-HIV infected and were 748 HIV-positive, in this cohort the mortality at 9-months was 35%, and the more common predictors for increased mortality were the higher stage of Medical Research Council and the lower CSF lymphocyte cell count (22).

The relationships among initial disease severity, long-term structural changes, and altered cognitive function in patients with TBM have not been fully studied, Chen Hsiu-Ling et al, in 2015 (23) corroborate that initial disease’s severity correlated to long-term structural abnormalities and cognitive function impairments. Individual disease severity assessed in acute illness is related to a long-term structural deficit, especially in the right caudate nucleus. These findings underscore the importance of evaluating initial disease severity to predict long-term disease outcomes. In our series only 37 (34.9%) had a cognitive evaluation.

An important fact to highlight is that the PanAmerican Health Organization reported in 2018 that the tuberculosis rate incidence in South America was 46.2 per 100,000 people, while in Mexico 25.9 per 100,000 respectively (24). Comorbidities are a challenge in our country, where 30-40% of tuberculosis of patients has diabetes; in our series only 15% reported this comorbidity.

The limitations of our study include the small sample of patients, the lack of availability of molecular diagnostic tools and the retrospective nature of the study itself. Despite of this our data shows important findings in a population of a referral neurological center.

## Conclusion

The clinical, laboratory, and neuroimaging characteristics between the population with HIV-positive and non-HIV infected shows no marked differences, particularly the presence of hydrocephalus. It is mandatory to start empirical treatment in all patients with a strong suspicion of TBM despite no mycobacterial isolation on CSF cultures or negative molecular diagnostic.

## Data Availability

the data are available upon request from the authors

## Declarations of interest

none

## Funding

This research did not receive any specific grant from funding agencies in the public, commercial, or not-for-profit sectors.

## Notes

### Competing Interest Statement

The authors have declared no competing interest.

### Author Declarations

This study protocol was approved (protocol 85/17) by the research and ethics committees of the Instituto Nacional de Neurologia y Neurocirugia Manuel Velasco Suarez

